# Fat fraction and iron concentration in lumbar vertebral bone marrow in the UK Biobank

**DOI:** 10.1101/2025.03.19.25324245

**Authors:** James R Parkinson, Marjola Thanaj, Nicolas Basty, Brandon Whitcher, E Louise Thomas, Jimmy D Bell

## Abstract

**Objectives:** This study aimed to assess the vertebrae bone marrow (VBM) fat and iron concentration in the UK Biobank imaging cohort (N = 26,531) using magnetic resonance imaging (MRI).

**Methods:** We measured the VBM fat using two approaches: fat fraction (FF) measured from Dixon MRI images and proton density fat fraction (PDFF) from multi-echo MRI scans, along with VBM iron concentration from multi-echo MRI images. We investigated sex-specific correlations between VBM measures and a range of anthropometric and lifestyle factors. Linear regression models were used to explore relationships between VBM measures, anthropometric and lifestyle factors, as well as disease status including osteoporosis and type-2 diabetes (T2D).

**Results:** VBM FF and PDFF were higher, while VBM iron concentration was lower in participants with osteoporosis and T2D (p < 0.00017). VBM FF and PDFF were positively associated with VAT, smoking, and T2D and were inversely associated with L1-L4 bone mineral density (BMD) and total skeletal muscle (p < 0.00017). VBM iron concentration was significantly positively associated with VAT, L1-L4 BMD, and alcohol intake.

**Discussion:** These findings enhance our understanding of VBM measures in metabolic health assessments, highlighting their role as potential indicators of metabolic health.

**Study importance:** *What is already known?:*

- Variations in bone marrow adipose tissue are linked to age, body composition, and clinical conditions such as type 2 diabetes, osteoporosis, and sarcopenia.
- Fat fraction (FF) derived from water-fat MRI is a robust method for assessing vertebral bone marrow (VBM) fat, which correlates with metabolic health markers.

*What does the study add?:*

- This study demonstrates sex-specific correlations of VBM fat fraction with age, body composition, and metabolic markers in the UK Biobank. It highlights relationships between VBM fat fraction and conditions such as sarcopenia, frailty, osteoporosis, type 2 diabetes, and back pain.
- This study identifies significant correlations between VBM iron concentration and anthropometric and disease variables, providing new insights into the role of iron deposition in bone health and metabolic processes.

*How might these results change the direction of research or the focus of clinical practice?:*

- The findings underscore the importance of including VBM fat fraction and iron concentration as imaging biomarkers in studies exploring metabolic and skeletal health.
- This study aims to shed light on sex-specific and condition-specific associations and may inform targeted interventions for metabolic and musculoskeletal conditions, especially in ageing populations, and encourage further research into the interplay between adiposity, bone health, and metabolic disorders.

## Introduction

Bone marrow adipose tissue accounts for approximately 8% of total body fat mass and plays a significant role in bone homeostasis, haemopoiesis and energy metabolism throughout the body (1). Elevated bone marrow fat content is observed in a wide range of conditions, including obesity, type 2 diabetes, osteoporosis and sarcopenia (2–4), thus understanding the pathophysiology and structural alterations that may accompany changes in the deposition of fat in the bone marrow is of great clinical importance which collectively contributes to rising healthcare costs and diminished quality of life (2, 5). Variations in bone marrow content are closely related to age and body composition (4), and its distinct physiology may be linked to the development of metabolic disorders (6). In contrast, white and brown adipose tissue depots and ectopic fat, bone marrow adiposity increases in states of caloric restriction, such as anorexia nervosa (7), and decreases upon weight recovery (8).

The structural composition of vertebral bone marrow (VBM) is predominantly assessed using chemical shift encoding-based water-fat magnetic resonance imaging (MRI) techniques, which permits the calculation of proton density fat fraction (PDFF) (9, 10). Assessment of bone marrow fat fraction has been employed in a number of smaller human cohort studies (<700 people), revealing associations with various aspects of metabolic health (11, 12). Bone marrow PDFF strongly correlates with visceral fat and insulin resistance, indicating the measurement may be used to screen subjects at risk of developing metabolic syndrome-associated features (13–15). Furthermore, the deposition of fat in VBM is associated with the arthritic inflammatory condition of joints and ligaments of the spine, ankylosing spondylitis, and may be considered a marker for early development of these conditions (16). Assessment of VBM fat fraction has also been shown to improve fracture discrimination power when used in combination with bone mineral density (BMD) (17, 18).

In addition to fat fraction, the MRI images allow measurement of organ iron deposition, which serves as an important marker of organ health. Iron overload in adipose tissue in models of obesity and insulin resistance (19), is linked to increased inflammation, decreased insulin sensitivity, and alterations in adipokine release and energy homeostasis (20). Furthermore, bone weakening, characterised by reduced bone mass, osteopenia, osteoporosis, and bone fractures, is a common feature in conditions associated with iron overload (21). In this study, we investigated lumbar marrow fat fraction and iron content in men and women and assessed variations with regard to anthropometric, lifestyle, indices of frailty, sarcopenia, osteoporosis and T2D in participants from the UK Biobank.

## Methods

### Data

The UK Biobank project is a population-scale, prospective cohort study of >500,000 participants aged 40-69 years recruited between 2006 and 2010 from across the UK (22), with a MRI scanning of the brain, cardiac and abdominal area in a sub-cohort of 100,000 participants. Participant data from the UK Biobank cohort was obtained through UK Biobank Access Application number 44584. The UK Biobank has approval from the North West Multi-Centre Research Ethics Committee (REC reference: 11/NW/0382). All methods were performed in accordance with the relevant guidelines and regulations, and informed consent was obtained from all participants. Researchers may apply to use the UK Biobank data resource by submitting a health-related research proposal that is in the public interest. More information may be found on the UKBB researchers and resource catalogue pages (https://www.ukbiobank.ac.uk).

### Phenotype Definitions

Anthropometric measurements, including age, weight (kg), height (cm), waist circumference (cm), and hip circumference (cm) were acquired at the UK Biobank imaging visit. From these values, BMI (kg/m^2^) and waist-to-hip ratio were calculated. Handgrip strength (HGS) was obtained from the self-reported dominant hand. If participants reported using both hands, the mean of the right and left hand was utilised. Participants with HGS <7kg were identified as outliers and excluded from further analysis. Ethnicity was defined based on the self-reported ethnic background at the initial assessment visit categorised as “White” and other ethnic background (due to small numbers of non-white participants in this dataset (3.1%)).

Following previously established quality control criteria, a metabolic equivalent task (MET) was taken at the UK Biobank imaging visit (23). Vigorous MET was used in our analysis as a more reliable indicator of high-intensity physical activity. All physical activity measures, alcohol intake frequency and smoking status were self-reported at the UK Biobank imaging visit.

The Townsend deprivation index is based on four variables: households without a car, overcrowded households, households not owner-occupied and persons unemployed and was taken at the initial assessment visit. Insulin-like growth factor 1 (IGF-1) was measured in nanomole/L at the initial assessment visit.

### Disease definitions

Sarcopenia was defined as low muscle quality as recommended by EWGSOP2 (24), which combines both low muscle mass with a HGS <27 kg in men and <16 kg in women and low muscle strength with dual X-ray absorptiometry (DXA)-measured appendicular lean mass (ALM)/height²: 5.5/7.0 kg/m² (female/male participants). Frailty was defined using the criteria adopted by Hanlon et al., (25) for use with self-reported UK Biobank questionnaire responses which require the presence of three out of five indicators including weight loss, exhaustion, no or only light physical activity in the last four weeks, slow walking speed, and low HGS (23). Additional classifications included pre-frailty (one or two of the five indicators), and not frail (none of the indicators). All frailty indicators were recorded during the UK Biobank imaging visit. Type-2 Diabetes (T2D) was defined based on the International Classification of Diseases 10th edition (ICD10) and self-reported codes for type-2 diabetes using E11, and 1220 or 1223, respectively (26). Osteoporosis was defined based on the ICD10 M80-M82 and based on WHO criteria to define osteoporosis (L1-L4, femoral neck and hip T-score ≤ −2.5) (11). Back pain was defined based on self-reported codes 6159 and 3571, as acute (if they had back pain for less than 3 months), and chronic back pain (if they had back pain for more than 3 months) (27).

### Image Analysis

Details of the UK Biobank MR abdominal protocol utilised in this study have previously been published (28, 29). The image-derived phenotypes (IDPs) included in this paper arise from the neck-to-knee Dixon MRI acquisition and separate single-slice multi-echo MRI acquisition for the Iterative Decomposition of Water and Fat with Echo Asymmetry and Least-Squares Estimation (IDEAL) liver. All MRI data were analysed using our dedicated image-processing pipelines for the neck-to-knee Dixon and single-slice multi-echo MR acquisitions, including the deep learning algorithms used to segment organs and tissue, as well as the estimation of PDFF and R2* values (29). Conversion from R2* to iron concentration followed McKay et al. (30). Our pipeline generates more than 30 IDPs from the neck-to-knee Dixon data, including VBM compartments from T1 (the first thoracic vertebra) to S1 (the first sacral vertebra). For training the deep learning segmentation model specifically on the VBM compartments, we utilised manual annotations from 120 participants. The Dice similarity coefficient on 20% out-of-sample test data was 0.83. The VBM segmentations were projected into the single-slice multi-echo data to obtain a two-dimensional region-of-interest (ROI) covering all voxels associated with VBM in the slice. VBM fat fraction (FF) was further computed to facilitate comparisons between FF and PDFF and to ensure consistency with previous studies computing FF in multi-site bone marrow (11, 31). The single-slice VBM segmentation was mapped to the Dixon FF maps, calculated as fat / (fat+water), by identifying its corresponding slice location within the full FF, enabling the quantification of the relative amount of water and fat within VBM tissue. Figure 1 illustrates the location of the single-slice multi-echo acquisition on the neck-to-knee Dixon scan, provides anatomical references of organs and tissue, and estimates of fat fraction and iron concentration in the vertebral bone marrow. Median values of FF were calculated from the VBM segmentation on the FF maps while median PDFF and iron concentration were computed from the VBM segmentation on the single-slice multi-echo liver acquisitions.

**Figure 1.**
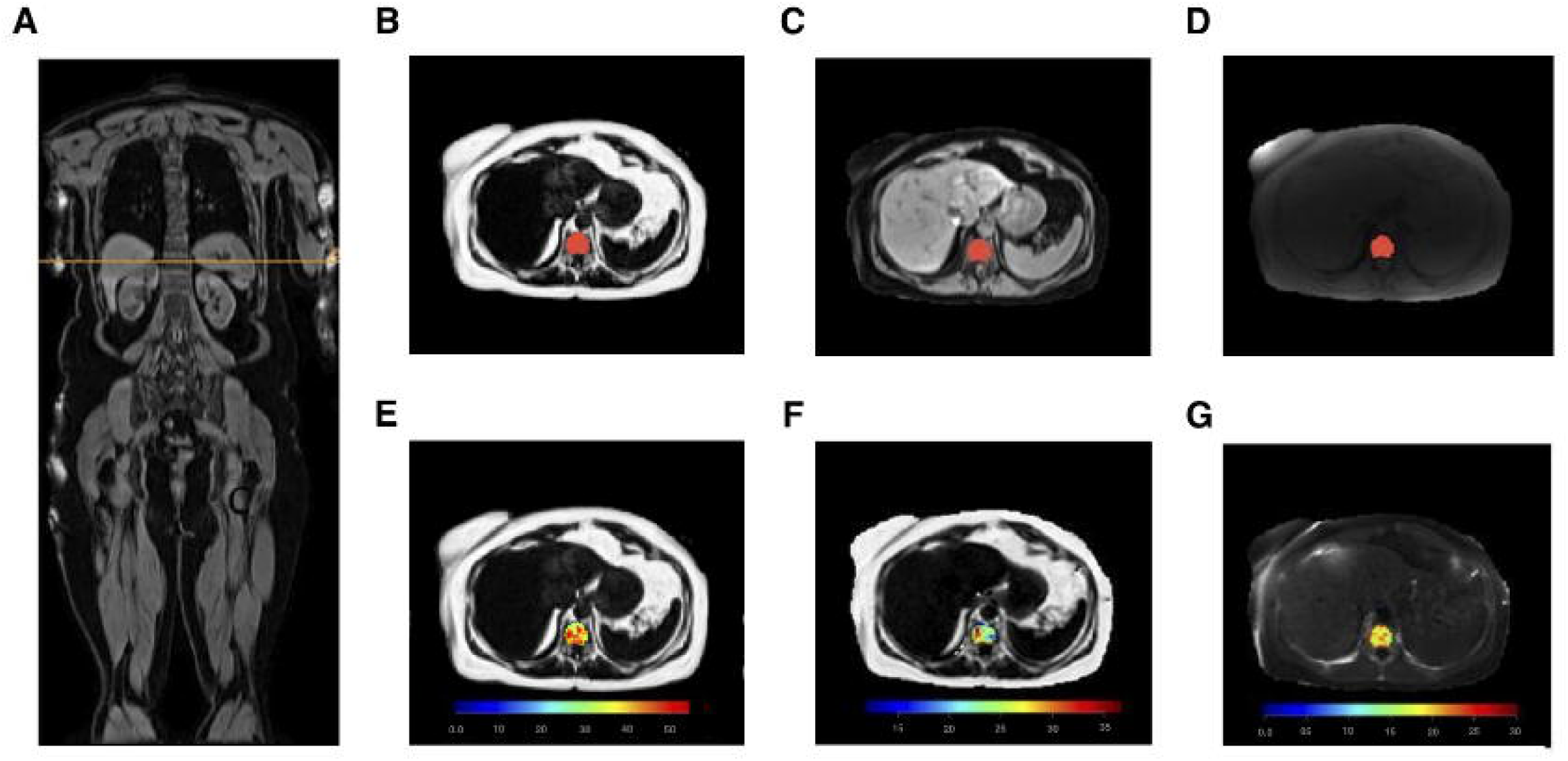
Localisation of vertebral bone marrow from reference MRI images. (A) Coronal slice of the neck-to-knee Dixon data showing the location of the single-slice multi-echo acquisition, (B) axial slice of the Dixon fat channel at the location with the VBM segmentation, (C) axial slice of the Dixon water channel at the location with the VBM segmentation, (D) the single-slice multi-echo data with the VBM segmentation, (E) voxelwise FF estimates (%) for the VBM segmentation, (F) voxelwise PDFF estimates (%) for the VBM segmentation, (G) voxelwise estimates of iron concentration (mg/g) for the VBM segmentation. Abbreviations: MRI, magnetic resonance imaging, PDFF, proton density fat fraction, VBM, vertebral bone marrow.

### Quality control

MRI data from 44,433 UK Biobank participants were screened for inclusion. Participants with missing single-slice multi-echo data and single-slice multi-echo data not intersecting with VBM segmentation (n=17,462), were excluded. Additionally, participants with segmentations of less than 64 voxels (1.9 cm^2^ in area) were excluded from the analysis (n=447). From the initial 44,433 participants, 26,524 were included in our analysis.

### Statistical analysis

All summary statistics, hypothesis tests, regression models and figures were performed using the R software environment for statistical computing and graphics (32). Descriptive statistics are expressed as mean and standard deviation in all tables and in the text. IDPs were tested for normality using the Kolmogorov-Smirnov test. The null hypothesis was rejected in all cases, and log transformations were applied to the median vertebral bone marrow iron concentration. While the null hypothesis was rejected for VBM FF and PDFF, there was limited deviation from normality when inspecting the quantile-quantile plot. The Wilcoxon rank-sum test was used to compare the means between two groups, while one-way ANOVA was used to compare multiple groups. Spearman’s rank correlation coefficient was used to assess monotonic trends between IDPs and variables of interest. we evaluated the impact of vertebral bone marrow FF, PDFF and iron concentration on anthropometric, lifestyle and disease factors using linear regression models, adjusting for age, ethnicity, height, vigorous MET, the Townsend deprivation index, IGF-1, L1-L4 BMD, alcohol, smoking, IDPs including total skeletal muscle volume, fat-to-muscle ratio in the mid-thighs (23) and VAT (29), as well as conditions including sarcopenia, frailty, osteoporosis, T2D and back pain. Summaries of the linear regression analysis are reported as regression coefficients (β) with standard errors in the tables and as standardised regression coefficients with 95% confidence intervals in the figure. The threshold for statistical significance of p-values was adjusted for the number of formal hypothesis tests performed. The Bonferroni-corrected threshold was 0.05/300 = 0.00017.

## Results

### Study Population

Baseline characteristics for the 26,531 participants included in the study are shown in Table 1. Of the 26,524 participants, 96.9% were of White ethnicity, and 51.0% were women, with an average age of 64.14 ± 7.52 years for women and 65.53 ± 7.77 years for men. The average BMI for women and men were 25.90 ± 4.68 kg/m^2^ and 26.89 ± 3.89 kg/m^2^ respectively. Within the study cohort, 3,458 participants were classified as having sarcopenia, 259 were classified as frail, 2,500 had osteoporosis, 1,403 were diagnosed with T2D, and 5,120 were diagnosed with back pain (of which 66.3% had chronic back pain).

**Table 1.**
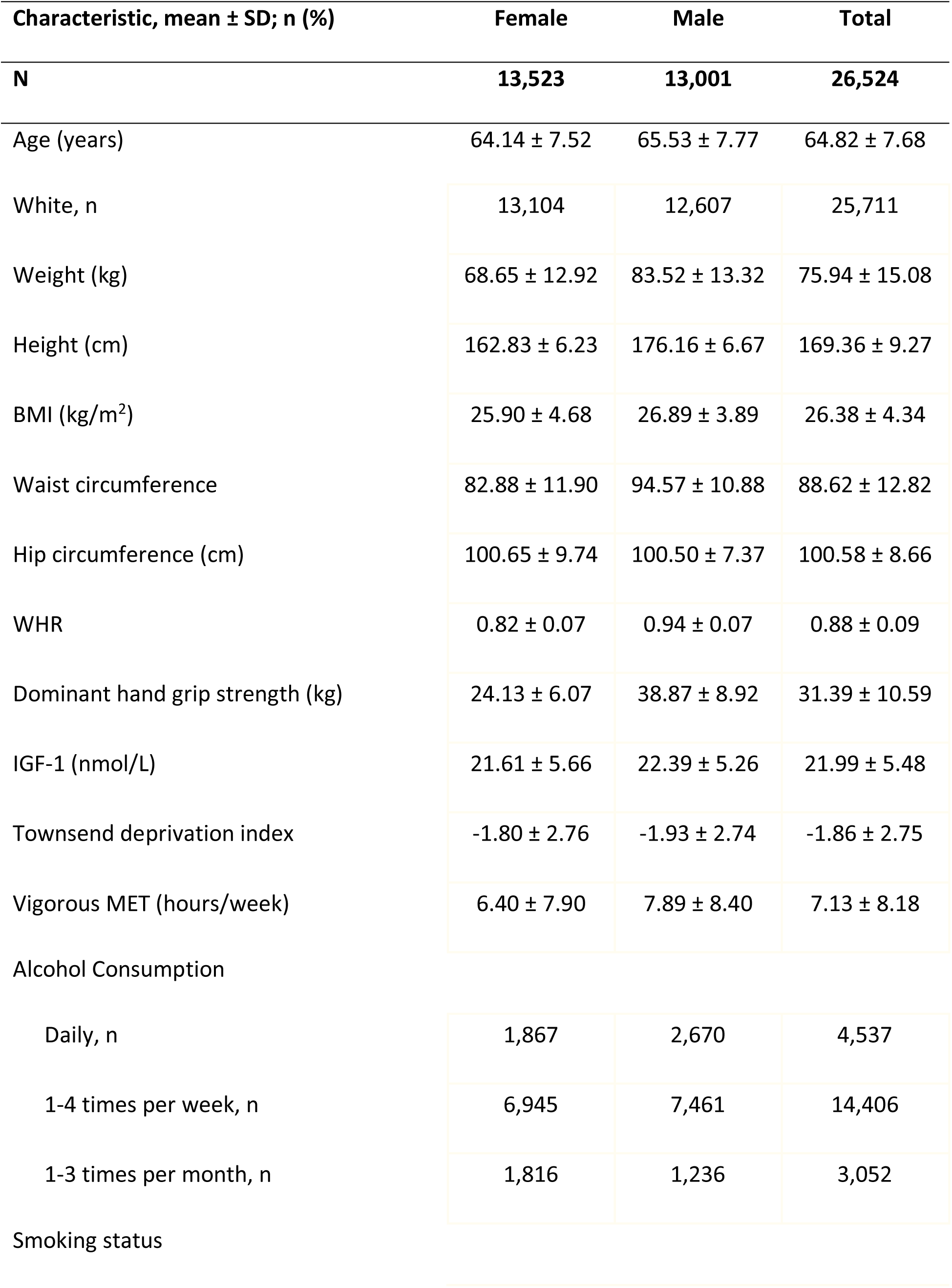
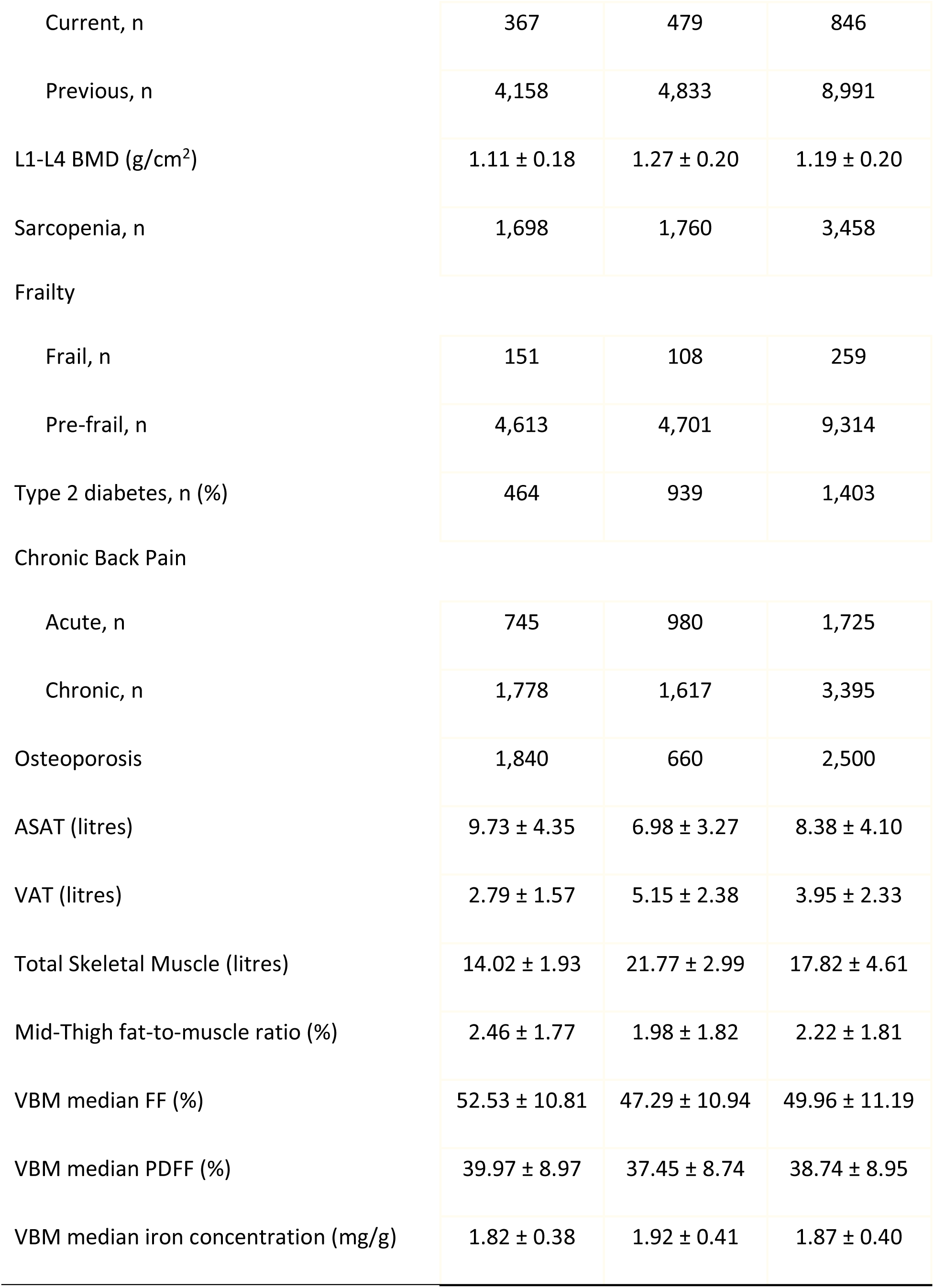
Demographics and participant characteristics. Abbreviations: ASAT, abdominal subcutaneous adipose tissue; BMD, bone mineral density; BMI, body mass index; IGF, insulin-like growth factor; FF, fat fraction; MET, metabolic equivalent task; PDFF, proton density fat fraction; VAT, visceral adipose tissue, VBM, vertebral bone marrow.

| Characteristic, mean ± SD; n (%) | Female | Male | Total |
| --- | --- | --- | --- |
| N | 13,523 | 13,001 | 26,524 |
| Age (years) | 64.14 ± 7.52 | 65.53 ± 7.77 | 64.82 ± 7.68 |
| White, n | 13,104 | 12,607 | 25,711 |
| Weight (kg) | 68.65 ± 12.92 | 83.52 ± 13.32 | 75.94 ± 15.08 |
| Height (cm) | 162.83 ± 6.23 | 176.16 ± 6.67 | 169.36 ± 9.27 |
| BMI (kg/m<sup>2</sup>) | 25.90 ± 4.68 | 26.89 ± 3.89 | 26.38 ± 4.34 |
| Waist circumference | 82.88 ± 11.90 | 94.57 ± 10.88 | 88.62 ± 12.82 |
| Hip circumference (cm) | 100.65 ± 9.74 | 100.50 ± 7.37 | 100.58 ± 8.66 |
| WHR | 0.82 ± 0.07 | 0.94 ± 0.07 | 0.88 ± 0.09 |
| Dominant hand grip strength (kg) | 24.13 ± 6.07 | 38.87 ± 8.92 | 31.39 ± 10.59 |
| IGF-1 (nmol/L) | 21.61 ± 5.66 | 22.39 ± 5.26 | 21.99 ± 5.48 |
| Townsend deprivation index | -1.80 ± 2.76 | -1.93 ± 2.74 | -1.86 ± 2.75 |
| Vigorous MET (hours/week) | 6.40 ± 7.90 | 7.89 ± 8.40 | 7.13 ± 8.18 |
| Alcohol Consumption |  |  |  |
| Daily, n | 1,867 | 2,670 | 4,537 |
| 1-4 times per week, n | 6,945 | 7,461 | 14,406 |
| 1-3 times per month, n | 1,816 | 1,236 | 3,052 |
| Smoking status |  |  |  |
| Current, n | 367 | 479 | 846 |
| Previous, n | 4,158 | 4,833 | 8,991 |
| L1-L4 BMD (g/cm<sup>2</sup>) | 1.11 ± 0.18 | 1.27 ± 0.20 | 1.19 ± 0.20 |
| Sarcopenia, n | 1,698 | 1,760 | 3,458 |
| Frailty |  |  |  |
| Frail, n | 151 | 108 | 259 |
| Pre-frail, n | 4,613 | 4,701 | 9,314 |
| Type 2 diabetes, n (%) | 464 | 939 | 1,403 |
| Chronic Back Pain |  |  |  |
| Acute, n | 745 | 980 | 1,725 |
| Chronic, n | 1,778 | 1,617 | 3,395 |
| Osteoporosis | 1,840 | 660 | 2,500 |
| ASAT (litres) | 9.73 ± 4.35 | 6.98 ± 3.27 | 8.38 ± 4.10 |
| VAT (litres) | 2.79 ± 1.57 | 5.15 ± 2.38 | 3.95 ± 2.33 |
| Total Skeletal Muscle (litres) | 14.02 ± 1.93 | 21.77 ± 2.99 | 17.82 ± 4.61 |
| Mid-Thigh fat-to-muscle ratio (%) | 2.46 ± 1.77 | 1.98 ± 1.82 | 2.22 ± 1.81 |
| VBM median FF (%) | 52.53 ± 10.81 | 47.29 ± 10.94 | 49.96 ± 11.19 |
| VBM median PDFF (%) | 39.97 ± 8.97 | 37.45 ± 8.74 | 38.74 ± 8.95 |
| VBM median iron concentration (mg/g) | 1.82 ± 0.38 | 1.92 ± 0.41 | 1.87 ± 0.40 |

As previous studies have primarily utilised FF to characterise bone marrow composition across multiple skeletal sites (11), we investigated both VBM FF and PDFF to maintain consistency with existing literature and to examine the extent to which these two measures capture sex-specific differences in VBM fat composition. We showed that median VBM FF was 52.53 ± 10.81 % in women and 47.29 ± 10.94 % in men, while median VBM PDFF was 39.97 ± 8.97 % in women and 37.45 ± 8.74 % in men whereas median iron concentration was 1.82 ± 0.38 mg/g in women and 1.92 ± 0.41 mg/g in men.

We further investigated the effect of disease on VBM measures (Supplementary Table S1-S5). We found statistically significant higher VBM FF and PDFF in participants with sarcopenia, osteoporosis and T2D (when compared to participants without sarcopenia, osteoporosis and T2D, respectively) in both men and women, while VBM FF was statistically significantly higher in men with frailty when compared to not-frail men. VBM iron concentration was statistically significantly higher in women with pre-frailty when compared to not-frail women, while it was lower in men with T2D and in both men and women with osteoporosis (when compared to non-T2D and non-osteoporosis, respectively) (p < 0.00017). We found no difference in the VBM measures between participants with and without chronic (Supplementary Table S5) or acute (results not shown) back pain.

### Correlation for VBM FF, PDFF and Iron Concentration

The sex-specific correlation coefficients between variables of interest and median VBM PDFF and iron concentration are shown in Figure 2, Supplementary Figure S1. In summary, both VBM FF and PDFF were significantly positively correlated with age in both men and women (r = 0.29 for FF, r = 0.30 for PDFF in women; r = 0.24 for both FF and PDFF in men, p < 0.00017). There was also a weak but significant correlation with VAT, mid-thigh fat-to-muscle ratio, ASAT and BMI in both men and women, showing similar results with both VBM FF and PDFF (r < 0.2, p < 0.00017). There was an inverse correlation between VBM PDFF and total skeletal muscle (r = -0.13 in women; r = -0.15 in men, p < 0.00017) with stronger correlations shown with VBM FF (r = -0.17 in women; r = -0.20 in men, p < 0.00017). There were weaker but significant negative correlations with IGF-1, vigorous MET and HGS (r > -0.1, p < 0.00017) with both VBM FF and PDFF. Both VBM FF and PDFF were significantly negatively correlated with L1-L4 BMD in both men and women (r = -0.16 for FF, r = -0.14 for PDFF in women; r = -0.07 for FF, r = -0.05 for PDFF in men, p < 0.00017). All significant correlations with median VBM iron concentration were much weaker for all significant correlations, with the highest correlations shown for age, BMI, VAT and L1-L2 BMD (r ≥ -0.1, p < 0.00017).

**Figure 2.**
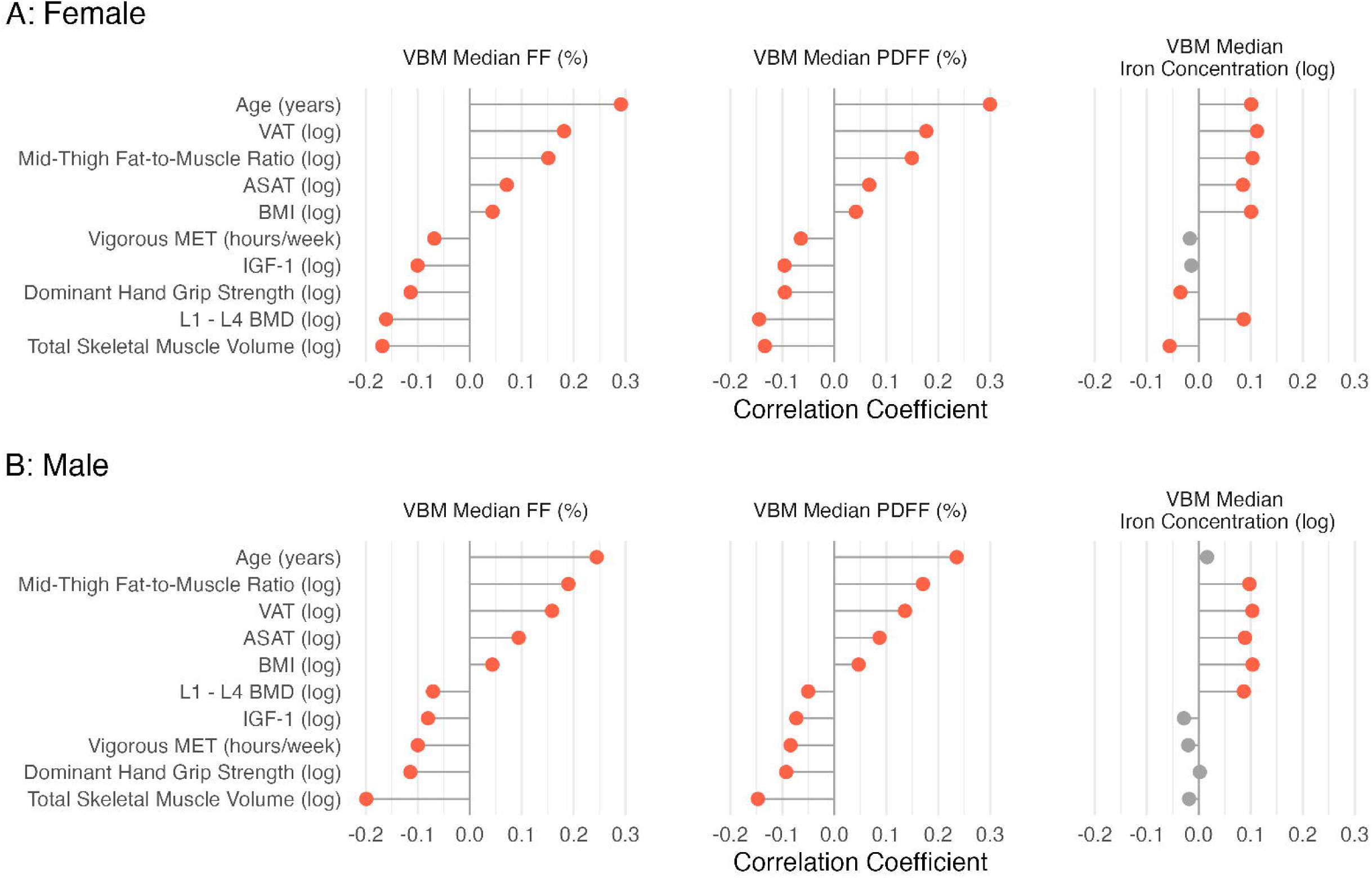
Correlation coefficients between variables of interest and median vertebral bone marrow FF, PDFF and iron concentration in female, male participants Correlation coefficients in red are statistically significant following correcting for multiple comparisons. IDP, image-derived phenotypes, FF, fat-fraction; PDFF, proton density fat-fraction; VBM, vertebral bone marrow. Significant associations for p-value below the Bonferroni correction (p = 0.00017) are shown in red, and non-significant associations in grey. Abbreviations: ASAT, abdominal subcutaneous adipose tissue; BMD, bone mineral density; BMI, body mass index; IGF, insulin-like growth factor; MET, metabolic equivalent task; PDFF, proton density fat fraction; VAT, visceral adipose tissue; VBM, vertebral bone marrow.

### Associations of VBM FF, PDFF and Iron with Anthropometric Characteristics and Disease

Linear regression models were used to investigate the effect of median VBM FF, PDFF and iron concentration on anthropometric, lifestyle factors and disease for men and women separately (Supplementary Table S6 and Figure 3). Median VBM FF and PDFF were significantly positively associated with age in both men and women and current smoking in women (p < 0.00017). VBM FF and PDFF were also positively associated with VAT (β = 3.66 %/log(L) for FF, β = 2.87 %/log(L) for PDFF in women and β = 3.50 %/log(L) for FF, β = 2.15 %/log(L) for PDFF in men p < 0.00017), while it showed a negative association with total skeletal muscle (β = -11.37 %/log(L) for FF, -8.04 %/log(L) for PDFF in women and β = -14.72 %/log(L) for FF, -8.36 %/log(L) for PDFF in men p < 0.00017). Median VBM FF and PDFF were significantly negatively associated with L1-L4 BMD in both men and women (β = -9.14 %/log(g/cm^2^) for FF, β = -7.13 %/log(g/cm^2^) for PDFF in women; β = -5.74 %/log(g/cm^2^) for FF, β = -4.05 %/log(g/cm^2^) for PDFF in men, p < 0.00017). Osteoporosis was positively associated with VBM FF in men (β = 2 %, p < 0.00017). A diagnosis of T2D was positively associated with VBM FF and PDFF, (β = 3.58 % for FF, β = 2.86 for PDFF in women and β = 2.65 % for FF, β = 1.94 for PDFF in men (p < 0.00017)).

**Figure 3.**
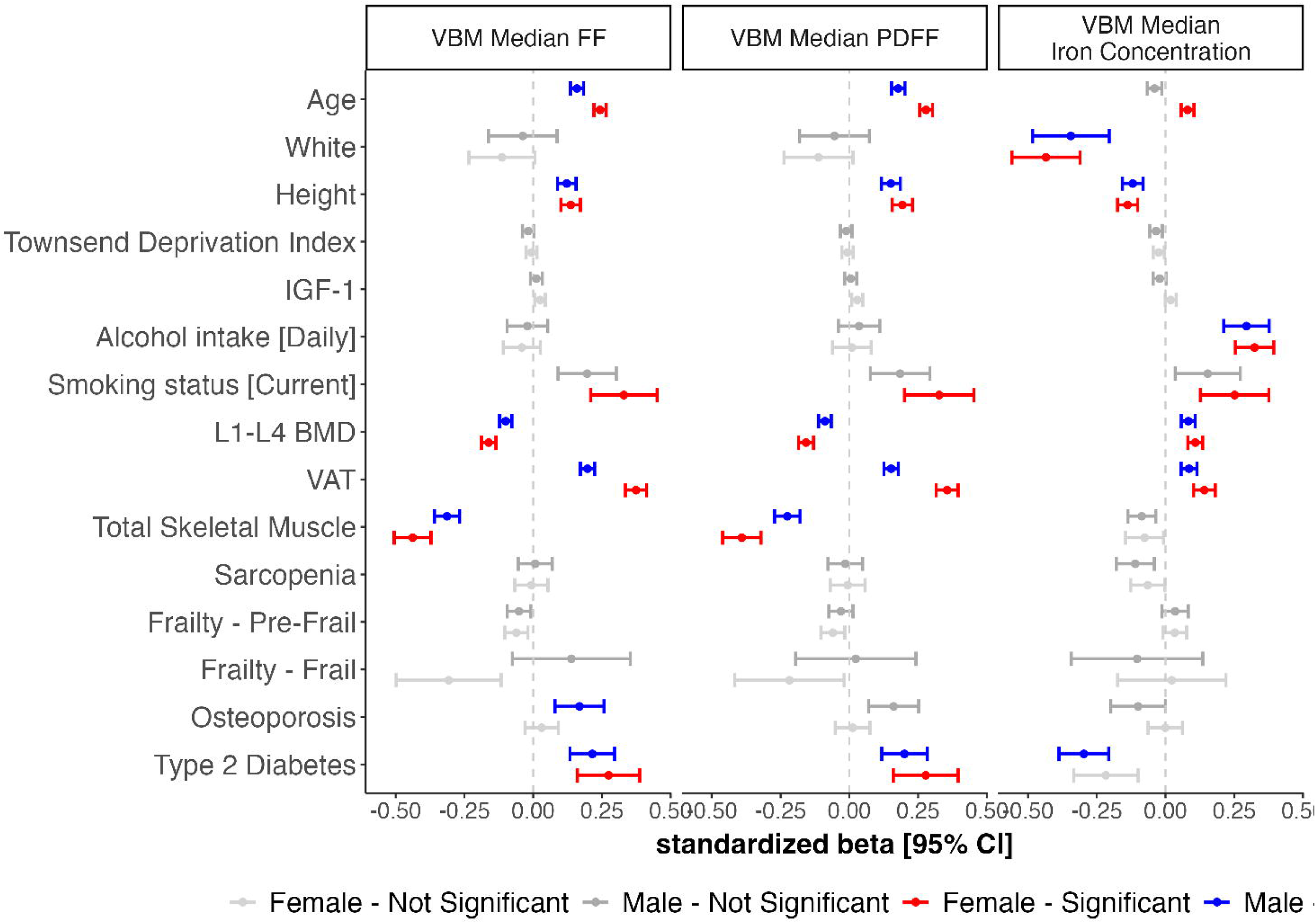
Summary of linear regression coefficients for median vertebral bone marrow FF, PDFF and iron concentration, representing the associations with the most relevant baseline characteristics on women and men separately. Standardised beta coefficients are displayed with 95% confidence intervals. Significant associations for p-value below the Bonferroni correction (p = 0.00017) are shown in red for women and blue for men, and non-significant associations in grey. Abbreviations: BMD, bone mineral density; IGF, insulin-like growth factor; FF, fat fraction;PDFF, proton density fat fraction; VAT, visceral adipose tissue; VBM, vertebral bone marrow.

Median VBM iron concentration was significantly positively associated with age and current smoking only in women and with daily alcohol intake, and L1-L4 BMD and VAT in both men and women (p < 0.00017), while there was a negative association with T2D in men (β = -0.06 log(mg/g) in men, p < 0.00017). We found no significant associations between VBM measures and sarcopenia, frailty or back pain. It is worth noting that R^2^ values were low, reflecting a poor fit in the linear regression models for VBM PDFF and iron concentration for both men and women, with the highest R^2^ values only observed for VBM FF and PDFF in women (adjusted R^2^ = 0.17 for both) (Supplementary Table S1-3).

## Discussion

In this study, we performed a comprehensive assessment of VBM FF, PDFF and iron concentrations in a large UK Biobank cohort. Our primary aim was to evaluate associations between VBM fat fraction, iron concentrations and anthropometric, as well as lifestyle characteristics, highlighting significant sex-specific associations that deepen our understanding of the variations in the regional fat accumulation, enabling greater insight into the anthropometric, lifestyle and disease factors. By examining fat fraction within VBM tissue and fat accumulation in VBM, measured as FF and PDFF, we aimed to ensure consistency with existing literature (11), while assessing the extent to which these metrics capture sex-specific associations in VBM fat composition.

We found that VBM FF and PDFF were positively correlated with age and VAT, in both men and women. While previous studies also reported associations between increased VBM PDFF and VAT, as well as ASAT and BMI (14), in our study, we found only weak correlations with BMI and ASAT (r ≤ 0.1). We further found an inverse correlation between VBM fat fraction (both FF and PDFF) and total skeletal muscle, IGF-1, vigorous physical activity, and hand grip strength, where ageing was associated with increased bone marrow adiposity, declining muscle function and physical activity, aligning with previous reports on vertebrae bone marrow fat (14, 33–35). Previous studies have also found positive associations between VBM fat and VAT and an inverse association with IGF-1 (14), supporting the role of IGF-1 as an important regulator of the fat and bone lineage, however, our linear regression models revealed significant associations with VAT but not with IGF-1.

Total muscle mass was significantly associated with reduced VBM FF and PDFF, whereas no significant association was found with sarcopenia. Our previous findings suggest that total muscle volume is more informative for sarcopenia, whereas muscle fat is more indicative of frailty (23). Nevertheless, we did not observe significant associations with frailty in our current models, however we showed that in men with frailty, VBM FF was higher when compared to not-frail men. Our results show that VBM fat accumulation is associated with reduced muscle volume, however previous studies suggest that muscle mass and quality loss is generally influenced by multifactorial factors, including environmental conditions, diseases, inflammation, and mitochondrial function (36, 37). Furthermore, despite previous studies reporting high L3 marrow fat in patients with back pain (38), we found no significant associations with back pain, highlighting the importance of further research in this area in order to provide us a better understanding of the impact of the fat accumulation in spinal bone marrow on back pain.

We also found an inverse association between lumbar BMD and both VBM FF and PDFF. Our results align with recent findings using deep learning techniques to facilitate multi-site bone marrow FF analysis in the UK Biobank, revealing site- and sex-specific characteristics that are crucial for understanding metabolic risk phenotypes (11, 31). These studies also reported significant differences in spine BMFF between osteoporosis and control groups primarily in women (11), while we identified significant differences in VBM FF and PDFF in both men and women with osteoporosis. Additionally, VBM PDFF was positively associated with T2D, aligning with previous findings suggesting that bone marrow fat may serve as a biomarker for glycemic control and potentially for late diabetic complications (39).

Our results further demonstrate significantly positive association between VBM iron concentration and factors such as alcohol intake, smoking, and VAT, with notable sex-specific trends and was lower in participants with osteoporosis. Interestingly, VBM iron concentration was positively associated with age only in women, suggesting a potential sex-dependent regulation of iron homeostasis in bone marrow that may be influenced by ageing (40). Furthermore, VBM iron concentration was positively associated with lumbar BMD, which is consistent with studies showing that higher dietary iron intake is linked to greater BMD (41) and lower risk of osteoporosis (42). However, while dietary iron has been associated with improved bone density in some studies, others suggest that iron overload may reduce bone quality (43), indicating a complex relationship between iron levels and bone health. These findings underscore the need for further research to explore the complex relationship between iron homeostasis and bone fragility in greater depth.

This study benefits from several strengths, including its large sample size, balanced gender distribution, and the use of advanced deep learning segmentation models to accurately measure VBM FF, PDFF, and iron concentrations. However, it is crucial to recognise its limitations. The UK Biobank, a substantial cross-sectional study, may be influenced by selection bias as it represents a ‘healthier’ group compared to the broader UK population and excludes younger individuals and cases of severe disease. Moreover, as the UK Biobank population is predominantly White, future work is needed to investigate diverse ethnic population data. Another potential limitation of this study is that the blood markers used were reported approximately 9 years before the MRI acquisition. Finally, the relatively low R^2^ values suggest that other factors may contribute to VBM measures. For instance relevant chronic inflammatory diseases in the spine such as ankylosing spondylitis which was diagnosed only on 83 participants (0.3%). Relevant markers such as L1-L4 bone mineral density, were only available on 75% of the imaging cohort, which reduced the number of observations in our analysis. Additional cross-sectional as well as longitudinal measurements will be required to assess age-related changes in disease cohorts.

## Conclusion

Our findings contribute to the broader understanding of how metabolic risk phenotypes extend beyond traditional measures, incorporating novel indicators such as VBM measures into multi-faceted risk models that can be associated with anthropometric, lifestyle characteristics, and disease. Our data further underscore the relevance of ectopic fat depots, suggesting that VBM FF, PDFF and iron may serve as complementary markers for comprehensive assessments of metabolic health.

## Declarations

Fully anonymised images and participant metadata were obtained through UK Biobank Access Application number 44584. The UK Biobank has approval from the North West Multi-Centre Research Ethics Committee (REC reference: 11/NW/0382), and obtained written informed consent from all participants before the study. All methods were performed in accordance with the relevant guidelines and regulations as presented by the appropriate authorities, including the Declaration of Helsinki.

## Data Availability

Our research was conducted using UK Biobank data. Under the standard UK Biobank data sharing agreement, we (and other researchers) cannot directly share raw data obtained or derived from the UK Biobank. However, under this agreement, all of the data generated, and methodologies used in this paper are returned by us to the UK Biobank, where they will be fully available. Access is obtained directly from the UK Biobank to all bona fide researchers upon submitting a health-related research proposal to the UK Biobank https://www.ukbiobank.ac.uk.

## DISCLOSURE

None of the authors has a conflict of interest to disclose.

## AUTHOR CONTRIBUTIONS

J.D.B., E.L.T., J.R.P and M.T. conceived the study. J.D.B., B.W., E.L.T., J.R.P and M.T. designed the study. M.T., B.W., J.R.P and N.B implemented the methods. M.T. and B.W. performed the data and statistical analysis. E.L.T., B.W., M.T., J.D.B., J.R.P, and N.B. drafted the manuscript. All authors read and approved the manuscript.

