## Supplementary_Material for "Fat fraction and iron concentration in lumbar vertebral bone marrow in the UK Biobank"

### Supplementary Figures

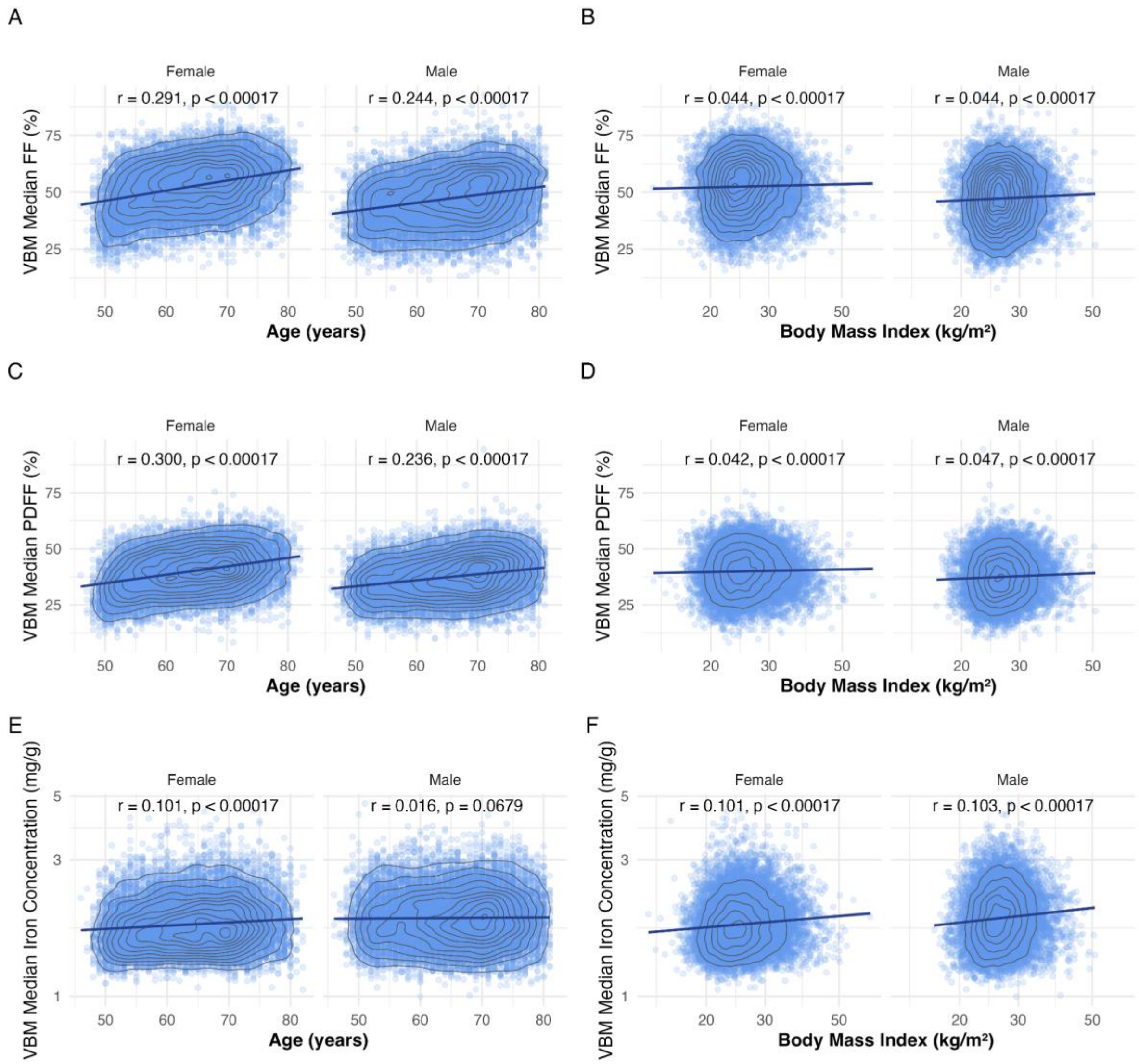


**Figure S1.** Gender specific distribution of vertebral bone marrow FF (**A**, **B**), PDFF (**C**, **D**) and iron concentration (**E**, **F**) by age (**A**, **C, E**), and BMI (**B**, **D, F**). BMI, body mass index; FF, fat-fraction; PDFF, proton density fat-fraction; VBM, vertebral bone marrow.

### Supplementary Tables

| Characteristic, mean ± SD; n (%) | Sarcopenia (1,698) | No Sarcopenia (11,501) | Sarcopenia (1,760) | No Sarcopenia (10,960) |
|---|---|---|---|---|
| | Female | | Male | |
| VBM median FF (%) | 54.07 ± 10.70 | 52.28 ± 10.81 ** | 49.70 ± 10.95 | 46.89 ± 10.89 ** |
| VBM median PDFF (%) | 41.10 ± 9.06 | 39.79 ± 8.94 ** | 39.01 ± 8.78 | 37.21 ± 8.71 ** |
| VBM median iron concentration (mg/g) | 1.82 ± 0.39 | 1.82 ± 0.38 | 1.89 ± 0.41 | 1.92 ± 0.42 * |

**Table S1.** Demographics and participant characteristics with and without sarcopenia. Significance refers to the p-value for a Wilcoxon rank-sums test, where the null hypothesis is the medians between the two groups (sarcopenia and no sarcopenia participants) being equal. * indicate statistically significant for $p < 0.05$, ** indicate statistically significant after Bonferroni correction ($p = 0.00017$). Note that there are overall 605 (324F/281M) missing data for sarcopenia. Abbreviations: FF, fat fraction; PDFF, proton density fat fraction; VBM, vertebral bone marrow.

| Characteristic, mean ± SD; n (%) | Frail[1] | Pre-Frail[2] | Not Frail[3] |
|---|---|---|---|
| Female | (151) | (4,613) | (7,708) |
| VBM median FF (%) | 52.81 ± 11.86 | 52.71 ± 10.84 | 52.24 ± 10.78 |
| VBM median PDFF (%) | 40.47 ± 9.64 | 40.04 ± 8.98 | 39.78 ± 8.93 |
| VBM median iron concentration (mg/g) | 1.82 ± 0.39 | 1.84 ± 0.39[3] | 1.81 ± 0.37[2] |

| Male | (108) | (4,701) | (7,334) |
|---|---|---|---|
| VBM median FF (%) | 51.17 ± 11.25[2] | 47.58 ± 11.12[1] | 46.95 ± 10.76 |
| VBM median PDFF (%) | 39.28 ± 8.84 | 37.74 ± 8.85 | 37.20 ± 8.63 |
| VBM median iron concentration (mg/g) | 1.85 ± 0.35 | 1.93 ± 0.42 | 1.91 ± 0.41 |

**Table S2.** Demographics and participant characteristics with and without osteoporosis. One-way ANOVA was used to compare the following groups: not frail - pre frail, not frail - frail and pre frail - frail participants. The numbers 1, 2, and 3 indicate those variables that were statistically significantly different between groups after Bonferroni correction (p = 0.00017). Note that there are overall 1,909 (1,051F/858M) missing data for dynapenia. Abbreviations: FF, fat fraction; PDFF, proton density fat fraction; VBM, vertebral bone marrow.

| Characteristic, mean ± SD; n (%) | Osteoporosis (1,840) | No Osteoporosis (11,683) | Osteoporosis (660) | No Osteoporosis (12,341) |
|---|---|---|---|---|
| | Female | | Male | |
| VBM median FF (%) | 55.56 ± 10.28 | 52.06 ± 10.81 ** | 50.59 ± 11.15 | 47.11 ± 10.90 ** |
| VBM median PDFF (%) | 42.25 ± 8.72 | 39.62 ± 8.95 ** | 39.78 ± 9.27 | 37.33 ± 8.70 ** |
| VBM median iron concentration (mg/g) | 1.78 ± 0.37 | 1.83 ± 0.38 ** | 1.83 ± 0.40 | 1.92 ± 0.42 ** |

**Table S3.** Demographics and participant characteristics with and without osteoporosis. Significance refers to the p-value for a Wilcoxon rank-sums test, where the null hypothesis is the medians between the two groups (osteoporosis and no osteoporosis participants) being equal. * indicate statistically significant for p < 0.05, ** indicate statistically significant after Bonferroni correction (p = 0.00017). Abbreviations: FF, fat fraction; PDFF, proton density fat fraction; VBM, vertebral bone marrow.

| **Characteristic, mean ± SD; n (%)** | **T2D (464)** | **No T2D (13,059)** | **T2D (939)** | **No T2D (12,062)** |
|---|---|---|---|---|
| | **Female** | | **Male** | |
| VBM median FF (%) | 56.86 ± 10.41 | 52.38 ± 10.79 ** | 51.26 ± 11.80 | 46.98 ± 11.81 ** |
| VBM median PDFF (%) | 43.42 ± 8.79 | 39.85 ± 8.95 ** | 40.16 ± 9.48 | 37.24 ± 8.65 ** |
| VBM median iron concentration (mg/g) | 1.80 ± 0.39 | 1.82 ± 0.38 * | 1.83 ± 0.38 | 1.93 ± 0.42 ** |

**Table S4.** Demographics and participant characteristics with and without T2D. Significance refers to the p-value for a Wilcoxon rank-sums test, where the null hypothesis is the medians between the two groups (T2D and no T2D participants) being equal. * indicate statistically significant for p < 0.05, ** indicate statistically significant after Bonferroni correction (p = 0.00017). Abbreviations: FF, fat fraction; PDFF, proton density fat fraction; T2D, Type-2 diabetes; VBM, vertebral bone marrow.

| **Characteristic, mean ± SD; n (%)** | **Back Pain (1,778)** | **No Back Pain (11,745)** | **Back Pain (1,617)** | **No Back Pain (11,384)** |
|---|---|---|---|---|
| | **Female** | | **Male** | |
| VBM median FF (%) | 53.34 ± 11.07 | 52.41 ± 10.76 * | 47.93 ± 11.16 | 47.20 ± 10.91 * |
| VBM median PDFF (%) | 40.56 ± 9.06 | 39.88 ± 8.95 * | 38.07 ± 8.89 | 37.36 ± 8.72 * |
| VBM median iron concentration (mg/g) | 1.81 ± 0.37 | 1.82 ± 0.38 | 1.91 ± 0.41 | 1.92 ± 0.42 |

**Table S5.** Demographics and participant characteristics with and without chronic back pain. Significance refers to the p-value for a Wilcoxon rank-sums test, where the null hypothesis is the medians between the two groups ( chronic back pain and no chronic back pain participants) being equal. * indicate statistically significant for p < 0.05, ** indicate statistically significant after Bonferroni correction (p = 0.00017). Abbreviations: FF, fat fraction; PDFF, proton density fat fraction; VBM, vertebral bone marrow.

| Characteristic | VBM Median FF (%) | | VBM Median PDFF (%) | | VBM Median Iron Concentration (log(mg/g)) | |
|---|---|---|---|---|---|---|
| | Female | Male | Female | Male | Female | Male |
| Age (years) | 0.357** | 0.239** | 0.326** | 0.211** | -0.001* | 0.002** |
| | -0.016 | -0.017 | -0.014 | -0.014 | -0.0003 | -0.0003 |
| White | -0.868 | -0.246 | -0.718 | -0.375 | -0.067** | -0.085** |
| | -0.672 | -0.698 | -0.559 | -0.569 | -0.014 | -0.013 |
| Height (cm) | 0.137** | 0.144** | 0.166** | 0.143** | -0.003** | -0.003** |
| | -0.021 | -0.02 | -0.018 | -0.017 | -0.0004 | -0.0004 |
| Vigorous MET (hours/week) | 0.004 | 0.001 | 0.004 | -0.007 | -0.00002 | 0.0003 |
| | -0.014 | -0.014 | -0.012 | -0.011 | -0.0003 | -0.0003 |
| Townsend Deprivation Index | -0.017 | -0.066 | -0.017 | -0.033 | -0.002* | -0.002* |
| | -0.04 | -0.042 | -0.033 | -0.035 | -0.001 | -0.001 |
| IGF-1 | 0.032 | 0.011 | 0.033* | 0.0005 | -0.001* | 0.001 |
| | -0.02 | -0.022 | -0.016 | -0.018 | -0.0004 | -0.0004 |
| Alcohol intake [Daily] | -0.611 | -0.372 | -0.029 | 0.231 | 0.057** | 0.066** |
| | -0.378 | -0.413 | -0.314 | -0.337 | -0.008 | -0.007 |
| Alcohol Frequency [1-4 times/week] | -0.433 | -0.107 | -0.191 | 0.309 | 0.036** | 0.033** |
| | -0.284 | -0.361 | -0.237 | -0.294 | -0.007 | -0.005 |
| Alcohol Frequency [1-3 times/month] | -0.165 | 0.402 | -0.161 | 0.394 | 0.011 | 0.015* |
| | -0.38 | -0.486 | -0.316 | -0.396 | -0.01 | -0.007 |
| Smoking status [Previous] | 0.660* | 0.671* | 0.547* | 0.672* | 0.008 | 0.011* |

| | | | | | | |
|---|---|---|---|---|---|---|
| | -0.236 | -0.238 | -0.197 | -0.194 | -0.005 | -0.004 |
| Smoking status [Current] | 3.670** | 2.144* | 2.897** | 1.621* | 0.033* | 0.052** |
| | -0.677 | -0.593 | -0.563 | -0.483 | -0.012 | -0.013 |
| L1-L4 BMD (nmol/L) | -9.140** | -5.737** | -7.131** | -4.053** | 0.082** | 0.109** |
| | -0.717 | -0.622 | -0.597 | -0.507 | -0.012 | -0.013 |
| VAT (log(L)) | 3.658** | 3.503** | 2.872** | 2.148** | 0.039** | 0.030** |
| | -0.188 | -0.232 | -0.156 | -0.189 | -0.005 | -0.004 |
| Total Skeletal Muscle (log(L)) | -11.365** | -14.719** | -8.041** | -8.363** | -0.071* | -0.028 |
| | -1.12 | -1.191 | -0.932 | -0.97 | -0.024 | -0.021 |
| Mid-Thigh Fat-to-Muscle Ratio (%) | -0.073 | 0.199* | -0.047 | 0.194* | 0.001 | 0.001 |
| | -0.071 | -0.081 | -0.059 | -0.066 | -0.002 | -0.001 |
| Sarcopenia | 0.19 | 0.108 | 0.155 | -0.113 | -0.019* | -0.01 |
| | -0.342 | -0.35 | -0.285 | -0.285 | -0.007 | -0.006 |
| Frailty - Pre-Frail | -0.620* | -0.565* | -0.491* | -0.269 | 0.006 | 0.007 |
| | -0.234 | -0.24 | -0.195 | -0.196 | -0.005 | -0.004 |
| Frailty - Frail | -3.180* | 1.84 | -1.760* | 0.382 | -0.017 | 0.003 |
| | -1.069 | -1.199 | -0.889 | -0.977 | -0.024 | -0.02 |
| Osteoporosis | 0.398 | 2.003** | 0.163 | 1.515* | -0.018 | -0.001 |
| | -0.34 | -0.5 | -0.283 | -0.408 | -0.01 | -0.006 |
| Type 2 Diabetes | 3.582** | 2.647** | 2.859** | 1.944** | -0.060* | -0.046** |
| | -0.632 | -0.452 | -0.526 | -0.369 | -0.009 | -0.012 |
| Back Pain - Chronic | 0.394 | 0.544 | 0.168 | 0.618* | -0.014* | -0.009 |
| | -0.322 | -0.342 | -0.268 | -0.278 | -0.007 | -0.006 |
| Constant | 53.329** | 47.065** | 40.555** | 37.102** | 0.665** | 0.628** |
| | -0.691 | -0.74 | -0.575 | -0.603 | -0.015 | -0.013 |
| AIC | 64294.4 | 63647.7 | 61108.6 | 60165.5 | -2996.1 | -4600.6 |

| | | | | | | |
|---|---|---|---|---|---|---|
| Observations | 8,669 | 8,510 | 8,669 | 8,510 | 8,510 | 8,669 |
| Adjusted $R^2$ | 0.174 | 0.124 | 0.169 | 0.099 | 0.044 | 0.065 |

**Table S6**. Summary of linear regression coefficients for median vertebral bone marrow FF, PDFF and iron concentration representing the associations with relevant baseline characteristics on women and men separately. Standard errors are shown in parentheses. An asterisk (*) indicates statistically significant for p-value < 0.05, ** indicate statistically significant after Bonferroni correction (p = 0.00017). Variables that are logarithmically transformed prior to adding in the model are shown as log. Abbreviations: BMD, bone mineral density; IGF, insulin-like growth factor; FF, fat fraction; MET, metabolic equivalent task; PDFF, proton density fat fraction; T2D, Type-2 diabetes; VBM, vertebral bone marrow.
